# Associations of clinical characteristics and antiviral drugs with viral RNA clearance in patients with COVID-19 in Guangzhou, China: a retrospective cohort study

**DOI:** 10.1101/2020.04.09.20058941

**Authors:** Xudan Chen, Yuying Zhang, Baoyi Zhu, Jianwen Zeng, Wenxin Hong, Xi He, Jingfeng Chen, Haipeng Zheng, Shuang Qiu, Ying Deng, Juliana CN Chan, Jian Wang

## Abstract

**Background:** The novel coronavirus disease 2019 (COVID-19) characterized by respiratory symptoms has become a global pandemic although factors influencing viral RNA clearance remained unclear to inform optimal isolation period and treatment strategies.

**Methods:** In this retrospective study, we included patients with confirmed COVID-19 admitted to Guangzhou Eighth People’s Hospital from 20^th^ January 2020 to 15^th^ March 2020. The associations of clinical characteristics and treatment regimens on time to viral RNA clearance were analyzed.

**Results:** We examined 284 consecutive COVID-19 cases, accounting for 82% of confirmed cases in Guangzhou during this period. At the time of reporting (20^th^ March 2020), 276 (97.2%) had recovered and were discharged from hospital with a median hospital stay of 18 days (interquartile range [IQR]:13-24). Overall, 280 patients achieved viral RNA clearance with a median length of 12 days (IQR: 8-16) after onset of illness. Amongst them, 66.1% had viral RNA cleared within 14 days, and 89.3% within 21 days. Older age, severity of disease, time lag from illness onset to hospital admission, high body temperature, and corticosteroid use were associated with delayed clearance of viral RNA. None of the antiviral regimens (chloroquine, oseltamivir, arbidol, and lopinavir/ritonavir) improved viral RNA clearance. The use of lopinavir/ritonavir was associated with delayed clearance of viral RNA after adjusting for confounders.

**Conclusion:** In patients with COVID-19, isolation for a minimum of 21 days after onset of illness may be warranted, while the use of antiviral drugs does not enhance viral RNA clearance.

**Brief Summary:** Viral RNA was cleared in 89% of the COVID-19 patients within 21 days after illness onset. The use of antiviral drugs (chloroquine, oseltamivir, arbidol, and lopinavir/ritonavir) did not shorten viral RNA clearance, especially in non-serious cases.

## Introduction

From the end of 2019, a new type of pneumonia caused by a novel coronavirus (severe acute respiratory syndrome coronavirus 2, SARS-CoV-2, also known as COVID-19), first broke out in Wuhan, spread rapidly throughout China, and became a global pandemic [1-4]. It has infected more than 1200,000 people worldwide and caused more than 67,000 deaths [5], posing enormous challenges to global public health.

To date, several epidemiological studies have reported the clinical characteristics of COVID-19 [1-3, 6-9]. Most of these studies with large sample size came from Wuhan, the center of the outbreak where sudden surge in demands for medical care has influenced timely treatment resulting in serious illness. Studies on COVID-19 patients in low incidence areas may provide additional insights into the clinical course of these patients.

The viral RNA dose determines a patient’s infectivity and is a key criterion in determining period of isolation and time of discharge from hospital [10-12]. There are no antiviral drugs specifically targeting COVID-19. The use of antiviral drugs approved for other conditions is largely empirical in these patients. Some antiviral drugs, such as chloroquine, arbidol, and lopinavir/ritonavir, have been investigated in small-scale studies and are recommended as options of antiviral regimens in China and some countries [11, 13-17]. However, the efficacy of these antiviral drugs on the SARS-CoV-2 RNA clearance remains largely unknown.

Guangzhou is a densely-populated metropolitan city in Southern China. In 2003, Guangzhou was heavily hit by SARS and the lessons learned from SARS have provided some experiences for the management of COVID-19. From 20^th^ January 2020 to 15^th^ March 2020, 347 patients were confirmed to have COVID-19 in Guangzhou; the majority of whom (n=284) were admitted to Guangzhou Eighth People’s Hospital designated for infectious disease. At the time of reporting (20^th^ March 2020), 97% of patients have been discharged with undetectable viral RNA. In this study, we summarized the clinical characteristics and patterns of use of antiviral drugs and their relationships with viral RNA clearance in patients with COVID-19.

## Methods

### Study design and participants

This is a retrospective study of consecutive patients with COVID-19 who were admitted to Guangzhou Eighth People’s Hospital between 20^th^ January 2020 and 15^th^ March 2020. The hospital was responsible for admitting the majority of COVID-19 confirmed cases in Guangzhou area. Patients were admitted from outpatient clinics or transferred from other hospitals. All confirmed COVID-19 cases, even asymptomatic, were isolated until viral RNA test became negative. The study was approved by the ethics committee of the Eighth Hospital of Guangzhou and all patients gave written informed consent.

### Definition

COVID-19 infection was confirmed by positive SARS-CoV-2 RNA test in respiratory tract samples using real-time RT-PCR method according to the manufacturer protocol (Daan Gene Co. Ltd, Guangzhou, China). For symptomatic patients, illness onset date was defined as the date when the symptoms (eg, fever, cough) first appeared. For asymptomatic patients, the date of first positive RT-PCR test was defined as the illness onset date.

According to the Chinese COVID-19 guideline [11], the severity of COVID-19 infection was grouped into four categories: mild, moderate, serious, and critical illness. Patients with mild illness did not have chest computed tomography (CT) findings, while those with moderate illness had features of pneumonia on chest CT but the symptoms were mild. Serious illness was defined by presence of at least one of the following criteria: (i) breathing rate >30/min; (ii) pulse oximeter oxygen saturation (SaO_2_) <93% at rest; (iii) ratio of partial pressure of arterial oxygen (PaO_2_) to fraction of inspired oxygen (FiO_2_) <300mmHg (1mmHg=0.133kPa). Critical illness was defined by presence of at least one of the following criteria: (i) occurrence of respiratory failure requiring mechanical ventilation; (ii) shock; (iii) failure of other organs requiring treatment in the intensive care unit (ICU). In our study, a group of patients were asymptomatic throughout the whole clinical course, although some of them had features of pneumonia on chest CT. To distinguish them from other patients, we defined them as asymptomatic patients.

Patients fulfilling all of the following criteria can be discharged: i) afebrile for >3 days, ii) improved respiratory symptoms, iii) resolution of infiltration shadows on pulmonary imaging, and iv) clearance of viral RNA. The latter was defined if nuclear acid test was negative in at least two sequential respiratory tract samples collected with more than 24 h interval [11]. In our study, the viral RNA tests in respiratory tract samples were repeated every 2-3 days since admission, and the first negative test date of sequential negative tests was defined as the viral RNA cleared day.

### Data collection

The records of patients with COVID-19 infection were extracted from the hospital electronic medical system. Demographic, clinical, laboratory, imaging and treatment data were collected and reviewed by trained physicians.

### Statistical analysis

Statistical analysis was performed using the IBM SPSS Statistics 20.0. Continuous variables were expressed as median (interquartile range, IQR), and categorical variables were expressed as number (%). Nonparametric test for continuous data and ordinal data, and chi-test for categorical data were used for between-group comparisons. Kaplan-Meier curve with log-rank test was used to evaluate the impact of demographic and clinical factors on time to viral RNA clearance and hospital stay, and Cox regression were used for controlling confounding factors. Logistic regression was used to evaluate the effect of antiviral regimens on day-14 clearance rate. A p value <0.05 (2-tailed) was considered to be statistically significant.

## Results

### Baseline characteristics

Overall, 284 patients with COVID-19 infection were included in this study, accounting for 82% of patients in Guangzhou during the reporting period (Table 1). The median age was 48 years old (IQR: 33-62), and 6 patients were adolescents (age <16 years old). A total of 131 (46.1%) patients were male, and 28 (9.9%) patients were smokers. The median incubation period was 6 days (IQR: 4-10), and the median interval from illness onset to hospital admission was 4 days (IQR: 2-7). Among these patients, 172 (62.0%) had a history of exposure to area of epicenter (Hubei province) or close contacts with people from Hubei, and 104 (36.6%) patients had at least one comorbidity, including hypertension (19.0%), diabetes (7.0%), cardiovascular diseases (5.3%), and malignancy (1.8%).

**Table 1.** Baseline characteristics and treatment outcomes in patients with COVID-19.

|  | Total (n=284) | clearance time from illness onset * |  | P value |
| --- | --- | --- | --- | --- |
|  |  | ≤14 days<br>(n=185) | >14 days since<br>onset (n=95) |  |
| No. patients with viral RNA clearance | 280 (98.6) |  | - |  |
| Age, median (IQR) years | 48 (33-62) | 47 (33-61) | 54 (37-63) | **0.047** |
| <16 years | 6 (2.1) | 4 (2.2) | 2 (2.1) |  |
| 16-49 years | 142 (50.0) | 99 (53.5) | 40 (42.1) | 0.329 |
| 50-64 years | 91 (32.0) | 56 (30.3) | 35 (36.8) |  |
| ≥65 years | 45 (15.8) | 26 (14.4) | 18 (18.9) |  |
| Sex |  |  |  | 0.717 |
| Female | 153 (53.9) | 99 (53.5) | 53 (55.8) |  |
| Male | 131 (46.1) | 86 (46.5) | 42 (44.2) |  |
| Smoking |  |  |  | 0.721 |
| No | 256 (90.1) | 167 (90.3) | 87 (91.6) |  |
| Yes | 28 (9.9) | 18 (9.7) | 8 (8.4) |  |
| History of exposure to Hubei or close<br>contact with people from Hubei | 176 (62.0) | 117 (63.2) | 56 (58.9) | 0.484 |
| Estimated incubation period, days | 6 (4-10) | 7 (4-10) | 6 (3-8) | 0.083 |
| Time lag from illness onset to hospital<br>admission, days | 4 (2-7) | 3 (1-6) | 5 (2-8) | **<0.0001** |
| **Severity of disease** |  |  |  |  |
| Asymptomatic | 17 (6.0) | 15 (8.1) | 2 (2.1) |  |
| Mild | 22 (7.7) | 15 (8.1) | 7 (7.4) |  |
| Moderate | 199 (70.1) | 133 (71.9) | 64 (67.4) | **0.036** |
| Serious | 40 (14.1) | 21 (11.4) | 19 (20.0) |  |
| Critical | 6 (2.1) | 1 (0.5) | 3 (3.2) |  |
| **Comorbidities** | 104 (36.6) | 70 (37.8) | 33 (34.7) | 0.610 |
| Hypertension | 54 (19.0) | 38 (20.5) | 16 (16.8) | 0.458 |
| Diabetes | 20 (7.0) | 11 (5.9) | 9 (9.5) | 0.278 |
| Cardiovascular disease | 15 (5.3) | 11 (5.9) | 3 (3.2) | 0.469 |
| Cerebrovascular diseases | 7 (2.5) | 4 (2.2) | 3 (3.2) | 0.920 |
| Respiratory disease | 10 (3.5) | 9 (4.9) | 1 (1.1) | 0.104 |
| Chronic liver disease | 12 (4.2) | 6 (3.2) | 5 (5.3) | 0.517 |
| Renal disease | 4 (1.4) | 3 (1.6) | 1 (1.1) | 1.000 |
| Malignancy | 5 (1.8) | 3 (1.6) | 2 (2.1) | 1.000 |
| **Symptoms and signs** |  |  |  |  |
| Maximum temperature on admission, °C | 37.0 (36.7-37.8) | 37.0 (36.7-37.6) | 37.1 (36.6-38.2) | 0.372 |
| <37.5°C | 184 (64.8) | 129 (72.1) | 54 (57.4) |  |
| 37.5-37.9 °C | 32 (11.6) | 21 (11.7) | 11 (11.7) |  |
| 38.0-38.9 °C | 52 (18.8) | 24 (13.4) | 27 (28.7) | **0.020¶** |
| ≥39.0 °C | 9 (3.2) | 5 (2.8) | 2 (2.1) |  |
| Fever before admission | 185 (65.1) | 116 (62.7) | 65 (68.4) | 0.343 |
| Cough | 161 (59.6) | 99 (56.6) | 58 (63.7) | 0.260 |
| Expectoration | 76 (28.1) | 47 (26.9) | 27 (29.7) | 0.627 |
| Fatigue | 64 (23.7) | 40 (22.9) | 21 (23.1) | 0.968 |
| Muscle pain | 48 (17.8) | 27 (15.4) | 19 (21.1) | 0.247 |
| Sore throat | 35 (13.0) | 22 (12.6) | 13 (14.3) | 0.695 |
| Catarrh symptoms | 35 (13.0) | 22 (12.6) | 11 (12.1) | 0.910 |
| Chest pain | 3 (1.1) | 3 (1.7) | 0 (0.0) | 0.553 |
| Chest distress | 21 (7.8) | 13 (7.4) | 8 (8.8) | 0.696 |
| Breath shortness | 30 (11.1) | 15 (8.6) | 14 (15.4) | 0.091 |
| Diarrhea | 21 (7.8) | 8 (4.6) | 11 (12.1) | **0.024#** |
| Nausea | 14 (5.2) | 9 (5.1) | 5 (5.5) | 0.903 |
| Vomiting | 7 (2.6) | 4 (2.3) | 3 (3.3) | 0.932 |
| Abdominal pain | 3 (1.1) | 1 (0.6) | 1 (1.1) | 1.00 |
| Anorexia | 47 (17.4) | 29 (16.6) | 18 (19.8) | 0.530 |
| Headache | 32 (12.1) | 23 (13.4) | 8 (9.1) | 0.313 |
| **Laboratory findings** |  |  |  |  |
| White blood cell (WBC) count, ×10<sup>9</sup>/L | 5.19 (4.14-6.39) | 5.15 (4.07-6.39) | 5.27 (4.18-6.45) | 0.835 |
| WBC > 9.5 × 10<sup>9</sup>/L | 12 (4.2) | 6 (3.2) | 6 (6.3) | 0.373 |
| WBC < 3.5 × 10<sup>9</sup>/L | 57 (20.1) | 40 (21.6) | 16 (16.8) | 0.344 |
| Neutrophil count, ×10<sup>9</sup>/L | 2.99 (2.16-3.87) | 3.03 (2.17-3.85) | 2.92 (2.15-4.01) | 0.717 |
| Lymphocyte count, ×10<sup>9</sup>/L | 1.35 (1.0-1.94) | 1.37 (1.04-1.99) | 1.31 (1.00-1.74) | 0.249 |
| Lymphocyte > 3.2 × 10<sup>9</sup>/L | 10 (3.5) | 7 (3.8) | 3 (3.2) | 1.00 |
| Lymphocyte < 1.1 × 10<sup>9</sup>/L | 93 (32.7) | 59 (31.9) | 32 (33.7) | 0.866 |
| Platelet count, ×10<sup>9</sup>/L | 195 (159-245) | 196 (159-242) | 195 (159-253) | 0.765 |
| Platelet count < 125 × 10<sup>9</sup>/L | 17 (6.0) | 11 (5.9) | 5 (5.3) | 0.816 |
| Hemoglobin, g/dl | 13.8 (12.5-15.5) | 14.2 (12.6-15.7) | 13.6 (12.1-14.7) | **0.041#** |
| Hemoglobin < 12.5 g/dl | 43 (15.2) | 28 (15.2) | 15 (15.8) | 0.900 |
| Alanine aminotransferase (ALT), U/L | 21.8 (14.4-33.3) | 21.3 (14.3-33.8) | 22.8 (15.5-33.4) | 0.264 |
| ALT > 40 U/L | 44 (16.5) | 27 (15.8) | 17 (18.7) | 0.551 |
| Aspartate aminotransferase (AST), U/L | 18.7 (15.1-26.9) | 18.3 (14.8-24.4) | 19.6 (15.9-29.3) | 0.056 |
| AST > 35 U/L | 36 (13.6) | 20 (11.8) | 15 (16.5) | 0.286 |
| Creatine kinase (CK), U/L | 54 (36-88) | 53 (31-81) | 52 (40-98) | 0.161 |
| Procalcitonin, ng/ml | 0.04 (0.03-0.07) | 0.04 (0.03-0.06) | 0.05 (0.03-0.09) | 0.171 |
| Procalcitonin > 0.05 ng/dl | 105 (38.3) | 62 (34.6) | 40 (44.0) | 0.135 |
| Albumin, g/L | 39.1 (35.3-42.3) | 39.5 (36.3-42.6) | 38.4 (34.8-41.6) | 0.102 |
| Albumin <40 g/L | 162 (57.0) | 98 (53.0) | 62 (65.3) | **0.049#** |
| Total bilirubin, umol/L | 10.1 (6.9-15.2) | 10.2 (6.7-15.1) | 9.8 (7.1-15.2) | 0.686 |
| Creatine, umol/L | 62.4 (50.7-76.8) | 62.1 (50.6-75.6) | 63.3 (50.4-79.2) | 0.520 |
| Lactate dehydrogenase (LDH), U/L | 190 (155-257) | 188 (155-249) | 196 (156-263) | 0.838 |
| LDH > 250 U/L | 73 (25.7) | 46 (24.9) | 25 (26.3) | 0.792 |
| Lactic acid, mmol/L | 1.8 (1.4-2.2) | 1.8 (1.4-2.1) | 1.9 (1.4-2.2) | 0.183 |
| Lactic acid >2.2 mmol/L | 59 (20.8) | 34 (18.4) | 23 (24.2) | 0.322 |
| D-dimer, ug/L | 1110 (780-1705) | 1110 (775-1725) | 1110 (780-1580) | 0.793 |
| D dimer >1000 ug/L | 150 (57.9) | 95 (57.2) | 53 (59.6) | 0.621 |
| CD4% | 42 (35-47) | 41.9 (35.4-47.0) | 43.7 (35.0-47.0) | 0.793 |
| CD8% | 27 (20-31.4) | 26.0 (20.0-30.2) | 29.0 (21.5-32.0) | 0.205 |
| CD4/CD8 ratio | 1.58 (1.22-2.18) | 1.64 (1.25-2.20) | 1.48 (1.12-2.15) | 0.273 |
| **Treatment** |  |  |  |  |
| ICU care | 24 (8.5) | 12 (6.5) | 10 (10.5) | 0.234 |
| Oxygen therapy | 184 (64.8) | 114 (61.6) | 67 (70.5) | 0.140 |
| Mechanical ventilation | 4 (1.4) | 0 | 2 (2.1) | 0.220 |
| Antibiotics | 213 (75.0) | 133 (71.9) | 76 (80.0) | 0.140 |
| Ribavirin | 2 (0.7) | 2 (0.1) | 0 | 0.550 |
| Chloroquine | 25 (8.8) | 14 (7.6) | 10 (10.5) | 0.402 |
| Corticosteroid | 31 (10.9) | 10 (5.4) | 19 (20.0) | **<0.0001¶** |
| Immunoglobulin | 8 (3.2) | 2 (1.1) | 4 (4.2) | 0.202 |
| Oseltamivir | 36 (12.7) | 17 (9.2) | 18 (18.9) | **0.019¶** |
| Lopinavir /ritonavir | 78 (27.5) | 43 (23.2 ) | 34 (35.8) | **0.026¶** |
| Arbidol | 69 (24.3) | 43 (23.2) | 26 (27.4) | 0.448 |
| **Outcome** |  |  |  |  |
| Discharge with recovery | 276 (97.2) | 183 (98.9) | 91 (95.8) |  |
| Remain in hospital | 3 (1.1) | 0 | 3 (3.2) |  |
| Transfer to other hospital | 4 (1.4) | 1 (0.5) | 1 (1.1) | 0.084 |
| Dead | 1 (0.4) | 1 (0.5) | 0 |  |
| Hospital stay, days | 18.0 (13.0-24.0) | 17.0 (12.0-23.0) | 21.0 (15.0-28.0) | **<0.0001** |
| Time from illness onset to viral RNA clearance, days | 12.0 (8.0-16.0) | 10.0 (6.0-12.0) | 19.0 (16.0-26.0) | **<0.0001** |
| Time from admission to viral RNA clearance, days | 7.0 (4.0-11.0) | 5.0 (3.0-8.0) | 14.0 (9.0-19.0) | **<0.0001** |

Data were presented as number (%) for categorical variables, and median (interquartile range) for continuous variables.
\*Four patient did not have information on viral RNA clearance and were excluded in the between-group analysis.
¶ Remained significant ( $P < 0.05$ ) after adjustment for age, severity of disease, and time lag from illness onset to hospital admission.

Fever (65.1%) and cough (59.6%) were the most common symptoms. The majority (64.8%) of patients had normal body temperature at the time of hospital admission. The laboratory findings were described in Table 1. Over half of patients (57.9%) had elevated D-dimer levels, and 38.3% had elevated procalcitonin levels. According to the severity of clinical manifestation and chest imaging, 22 (7.7%), 199 (70.1%), 40 (14.1%), and 6 (2.1%) patients were categorized into mild, moderate, serious, and critical illness, respectively, and 17 patients (6.0%) were asymptomatic throughout the whole clinical course although 11 of them had a pneumonia CT image on admission. No significant differences were observed in disease severity by sex and smoking status. Elderly patients and those with comorbidity were more likely to have serious and critical illness (Table 2).

**Table 2.**
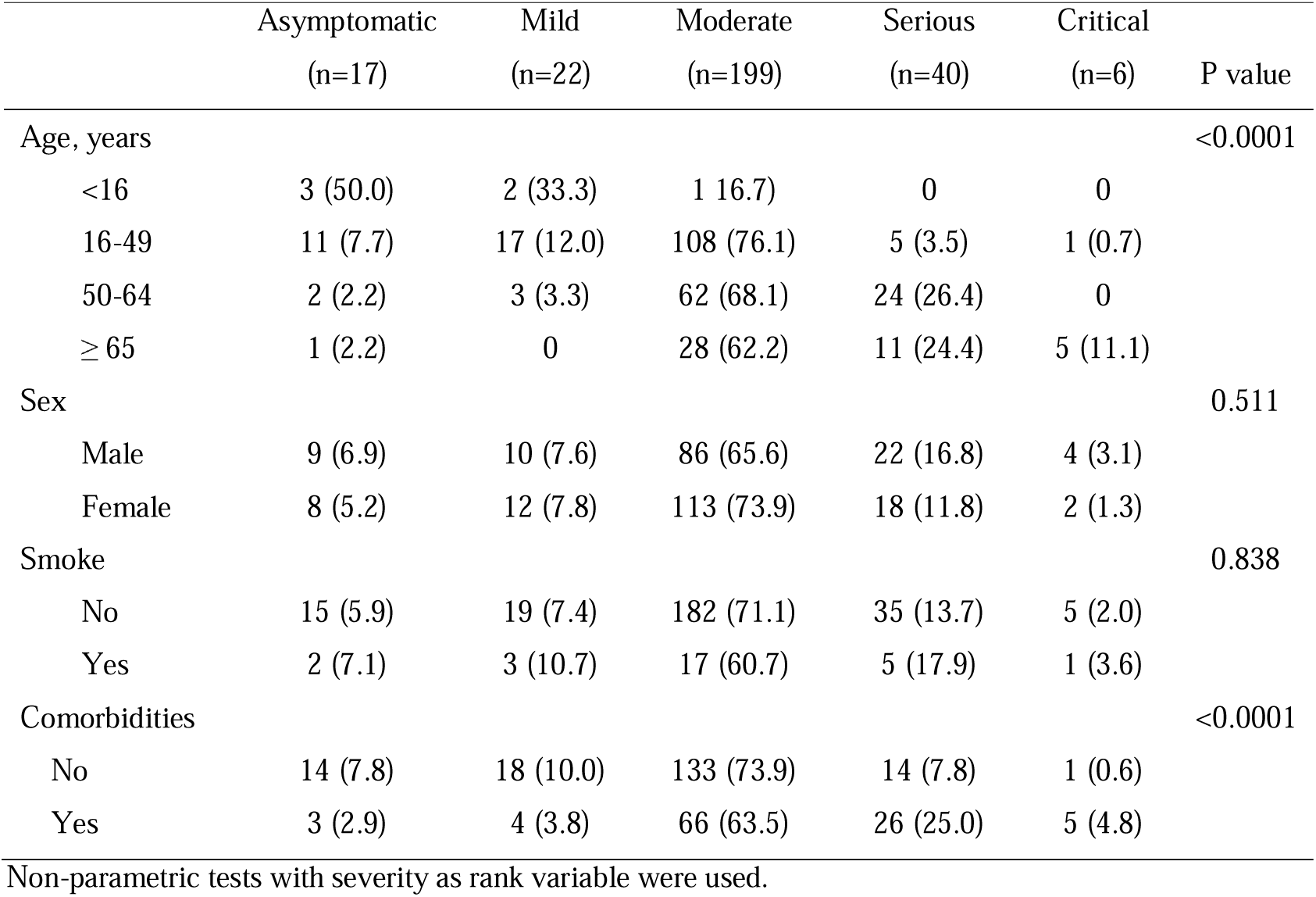
Distribution of COVID-19 severity by demographic and clinical characteristics.

### Treatment

The majority of patients received antibiotic treatment (75.0%) and oxygen therapy (64.8%). Four (1.4%) patients received mechanical ventilation, and 24 (8.5%) patients were admitted to ICU. A total of 25 (8.8%) patients received chloroquine, and 36 (12.7%), 78 (27.5%), 69 (24.3%) patients were treated with the antiviral drugs of oseltamivir, lopinavir/ritonavir, and arbidol, respectively. Thirty-one (10.9%) patients were treated with corticosteroid, 9 (3.2%) patients received immunoglobulin, and 2 (0.7%) patients received ribavirin (Table 1).

### Outcome and viral RNA clearance

At the time of reporting (20^th^ March 2020), 276 (97.2%) patients have recovered and were discharged from hospital with a median hospital stay of 18 days (IQR: 13-24, range: 6-46). One (0.4%) 81-year-old woman died 17 days after hospital admission. She had serious illness and comorbid pancreatic cancer, diabetes and cardiovascular disease at the time of admission. Four (three critical and one moderate) patients were transferred to other hospitals, and three (two moderate and one serious) patients were still recovering in the hospital.

A total of 280 (98.6%) patients with undetectable viral RNA had valid data on time to viral RNA clearance. The median duration from illness onset to viral RNA clearance was 12 days (IQR:8-16), and 7 days (IQR: 4-11) from hospital admission to clearance. Overall, 89.3% patients achieved viral RNA clearance within 21 days and 66.1% within 14 days (Figure 1)

**Figure 1.**
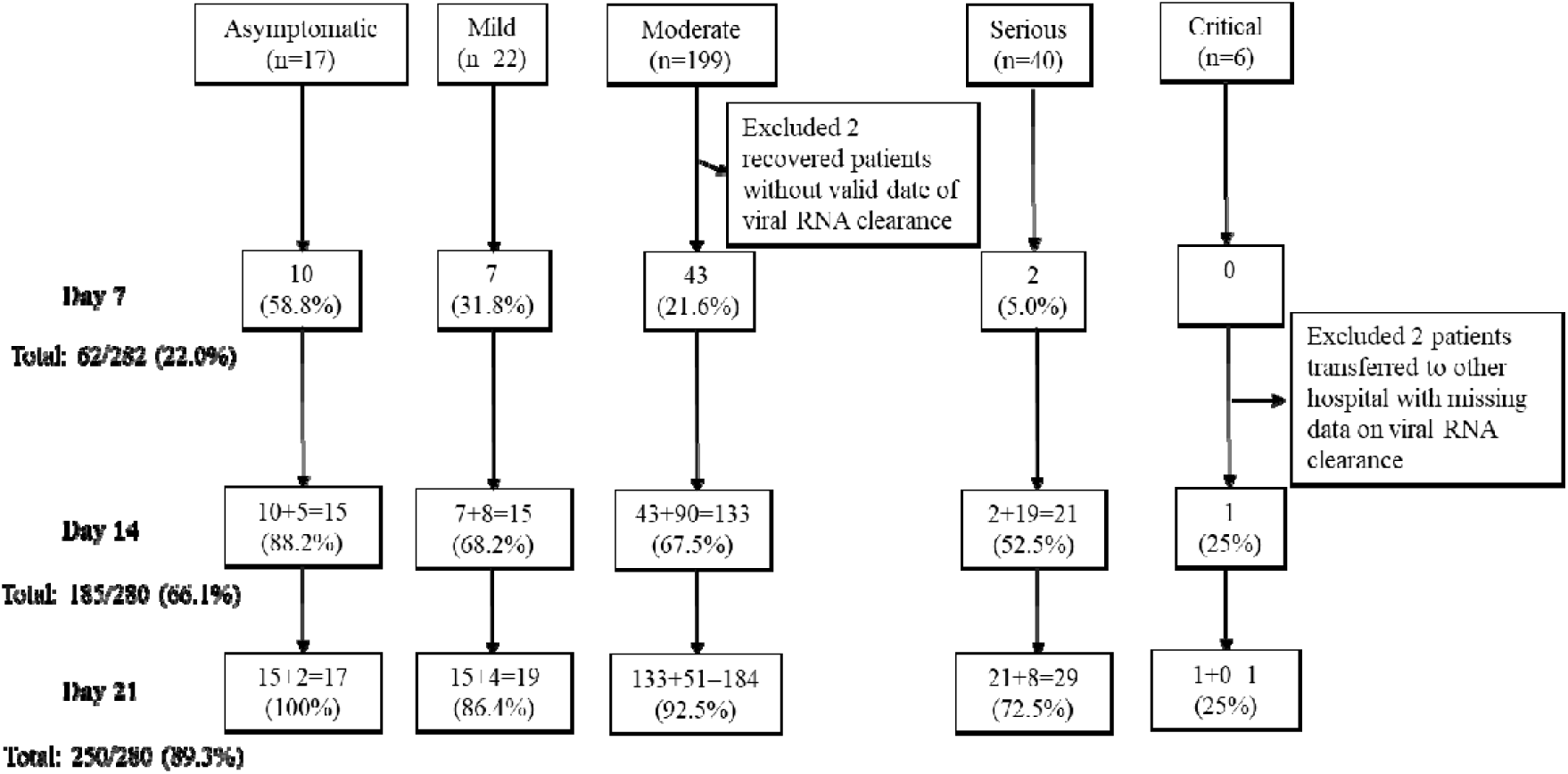
The cumulative viral RNA clearance rate at day 7, day 14, and day 21 by severity of COVID-19 infection. The diagram showed the cumulative clearance rate at day 7, day 14, and day 21 by severity of COVID-19 infection. Number (%) of patients were presented.

**Figure 2.**
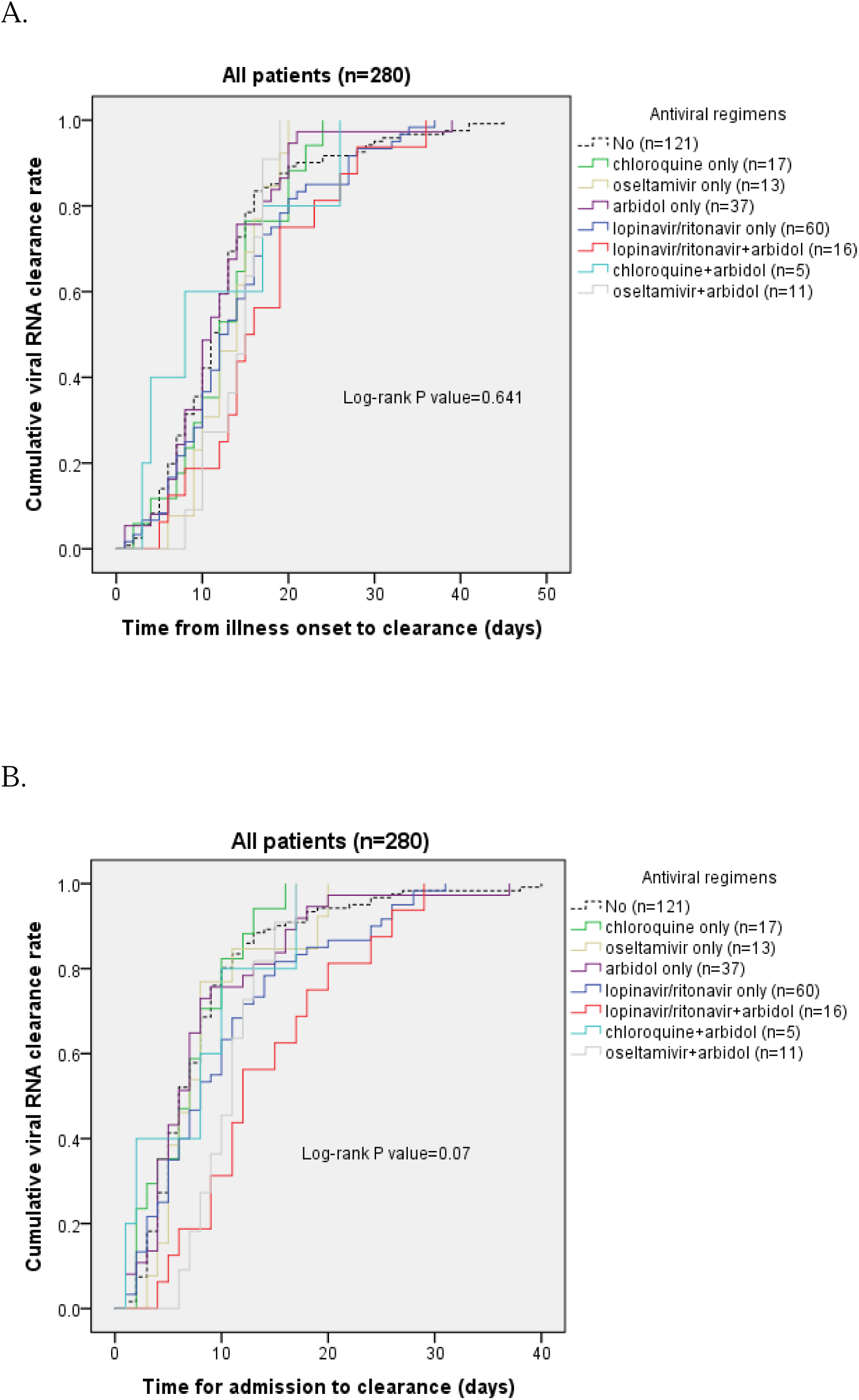
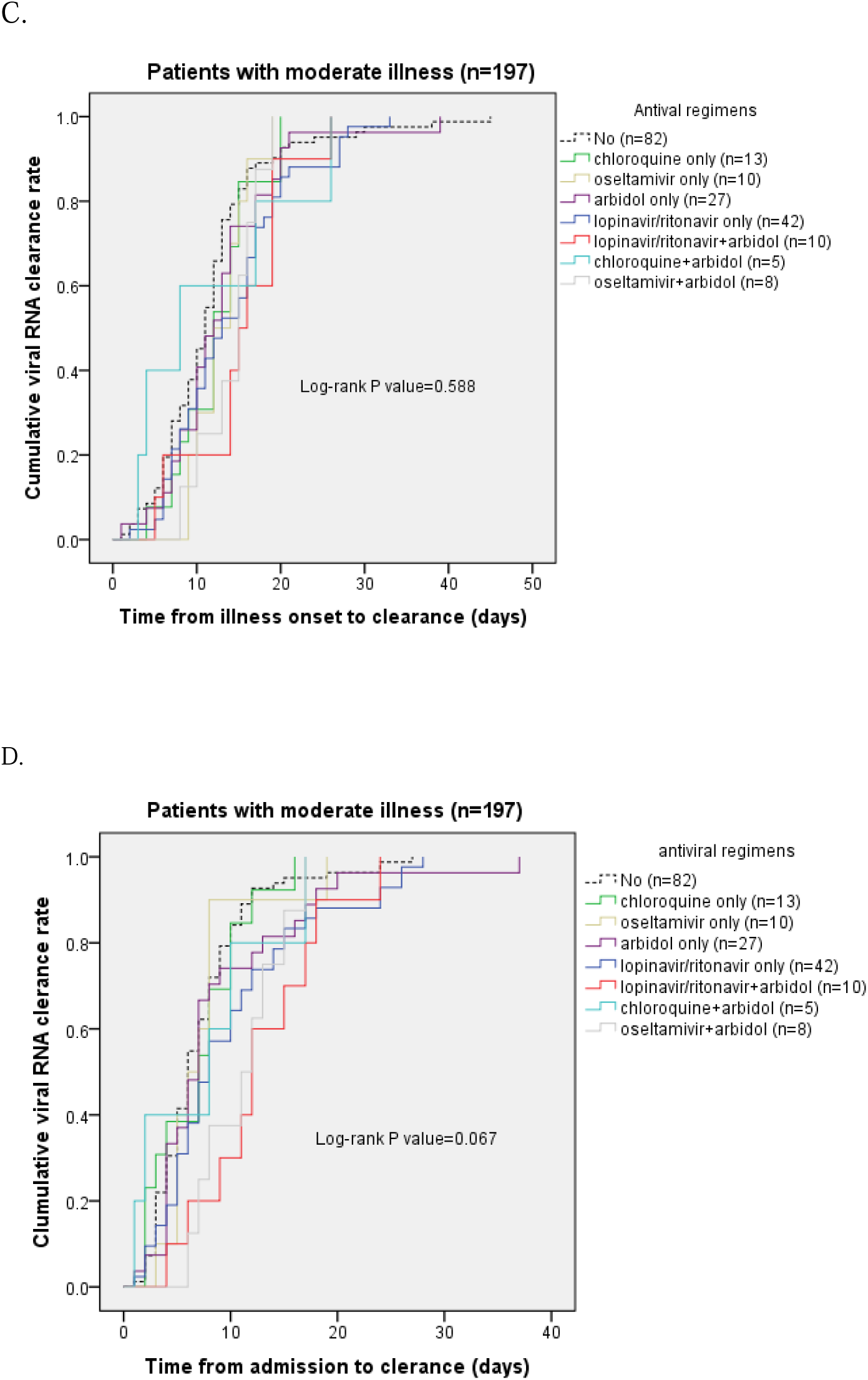
Kaplan-Meier curve of COVID-19 viral clearance rate by antiviral treatment regimens. (A) cumulative viral RNA clearance by time from illness onset in all patients with COVID-19 infection (n=280); (B) cumulative viral RNA clearance rate by time from hospital admission in all patients (n=280); (C) cumulative viral RNA clearance by time from illness onset in patients with moderate illness (n=204); (D) cumulative viral clearance by time from hospital admission in patients with moderate illness (n=204). Log-rank test was used for group comparisons.

### Factors and therapeutic regimens associated with viral RNA clearance

We compared the time to viral RNA clearance by demographic and clinical characteristics. As shown in Table 1 and Table 3, time lag from illness onset to hospital admission, old age, disease severity, high body temperature, and corticosteroid use were associated with delayed viral RNA clearance. Breath shortness, elevated white blood cell count, reduced serum albumin, oxygen therapy, and immunoglobulin use were also associated with delayed viral RNA clearance; however the association became insignificant after adjusting for age, disease severity and time lag from illness onset to hospital admission. No significant associations with sex, smoking status, comorbid diseases, and other disease symptoms were observed (Table 1 and Table 3).

**Table 3.**
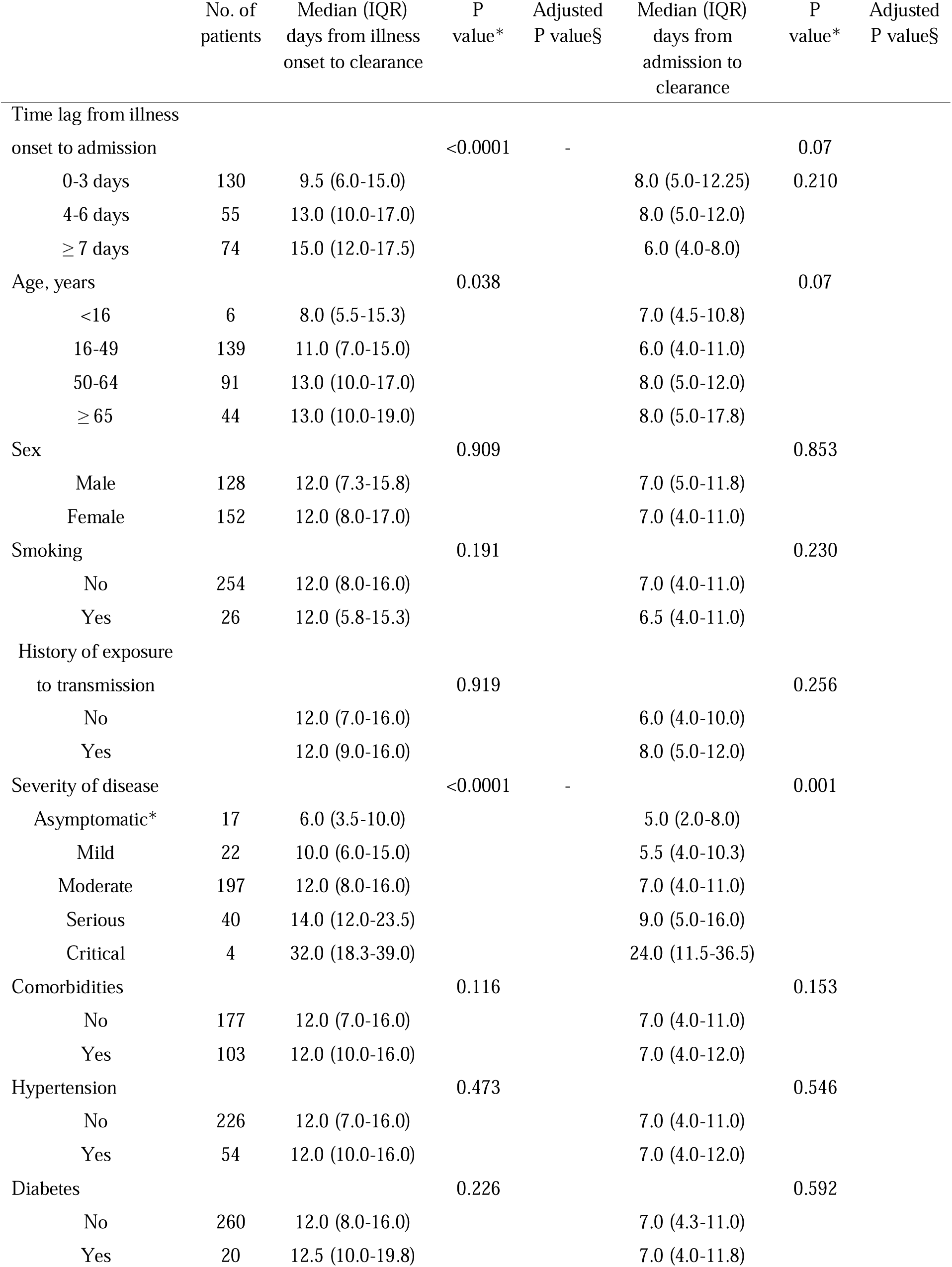
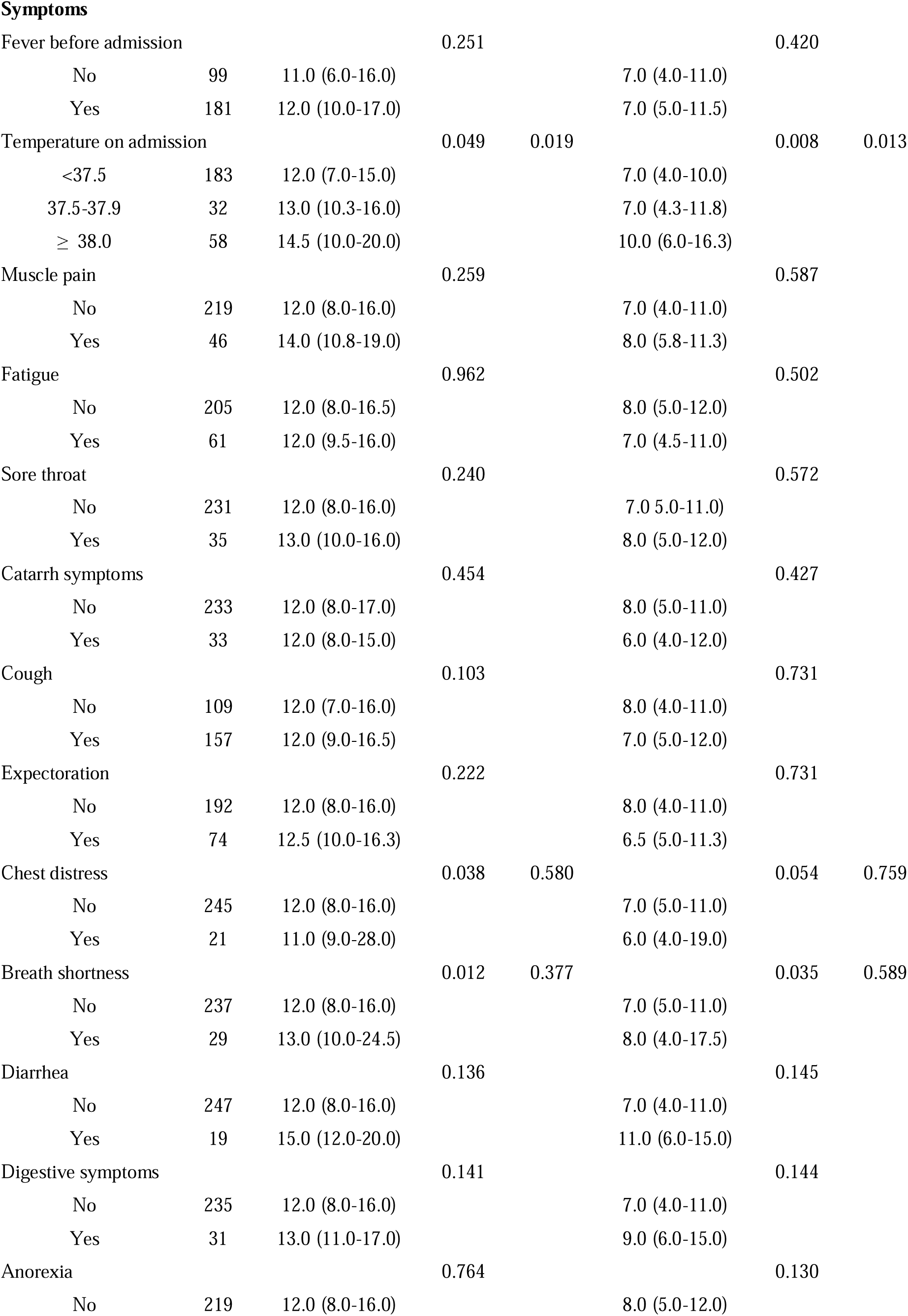
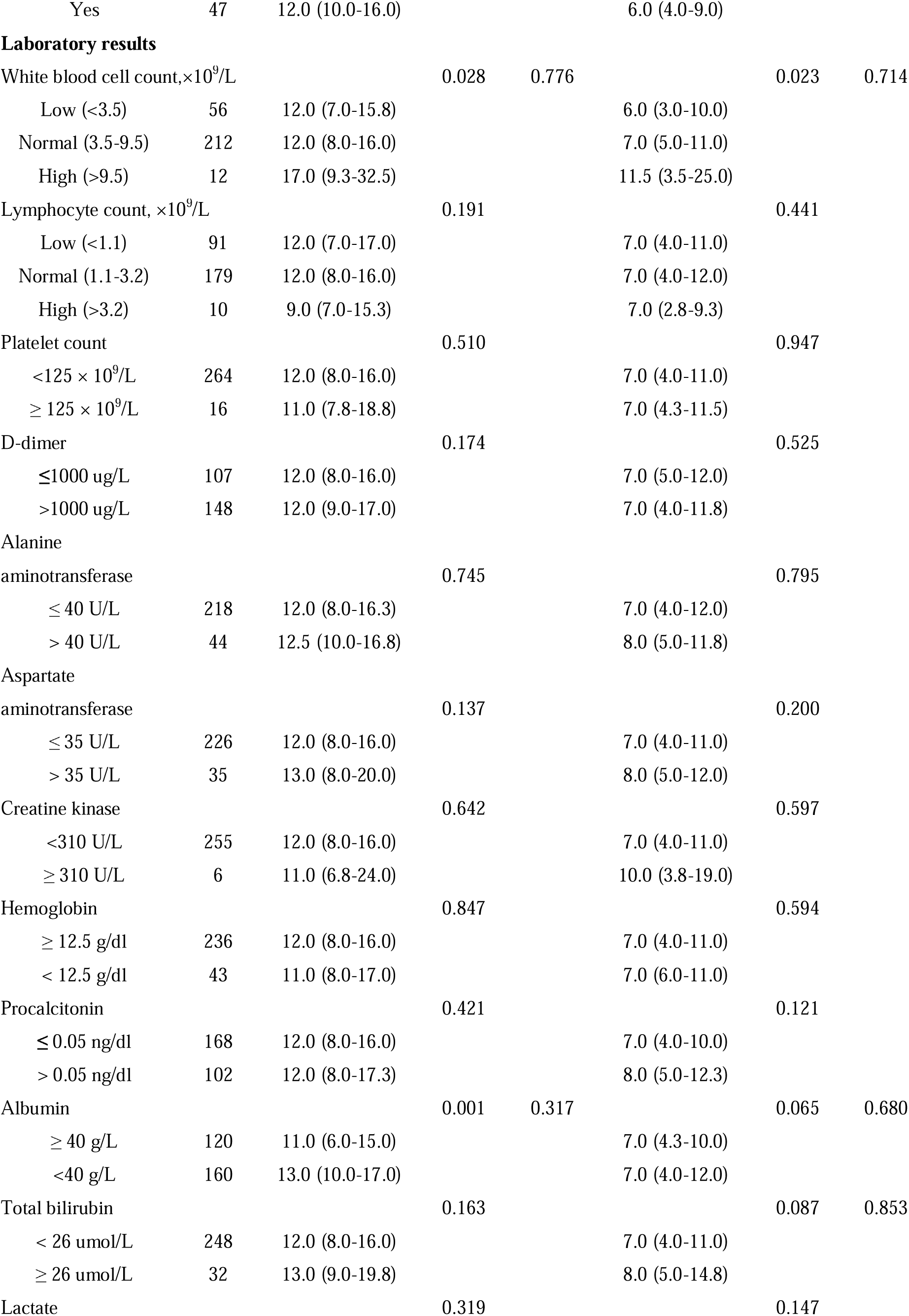
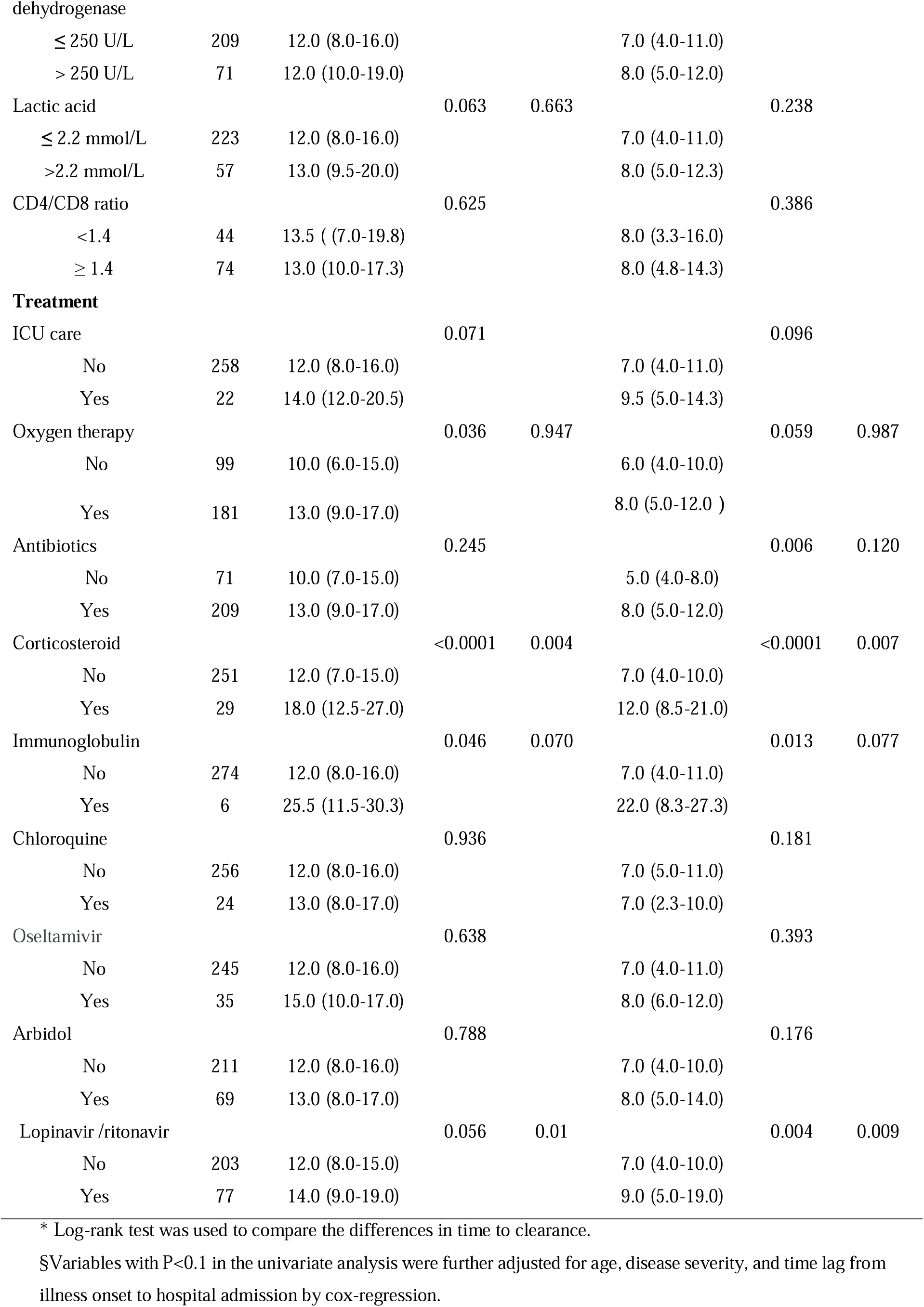
Viral detectable duration by demographic and clinical characteristics in patients with COVID-19.

There was no specific guideline on indication of use of antiviral drugs. We compared the effects of different antiviral drug combinations (chloroquine, oseltamivir, arbidol, and lopinavir/ritonavir) on viral RNA clearance. As shown in Table 4, 121 (43.2%) patients did not receive any antiviral drugs, and the median duration was 11 days (IQR: 7-15) from illness onset, and 6 days (IQR: 4-9) from hospital admission to viral RNA clearance. The use of antiviral drugs, as monotherapy or in combination, was not associated with improved clearance of viral RNA compared to those did not receive antiviral drugs. Patients treated with lopinavir/ritonavir, a combination therapy for treating HIV infection, tended to have delayed viral RNA clearance, even after controlling for age, disease severity, time lag from illness onset to hospital admission, temperature, and corticosteroid use. The combined use of two antiviral agents was also associated with delayed clearance of viral RNA after adjusting for confounders. The analysis focusing on patients with moderate illness yielded similar results (Table 4 and Figure 1).

**Table 4.**
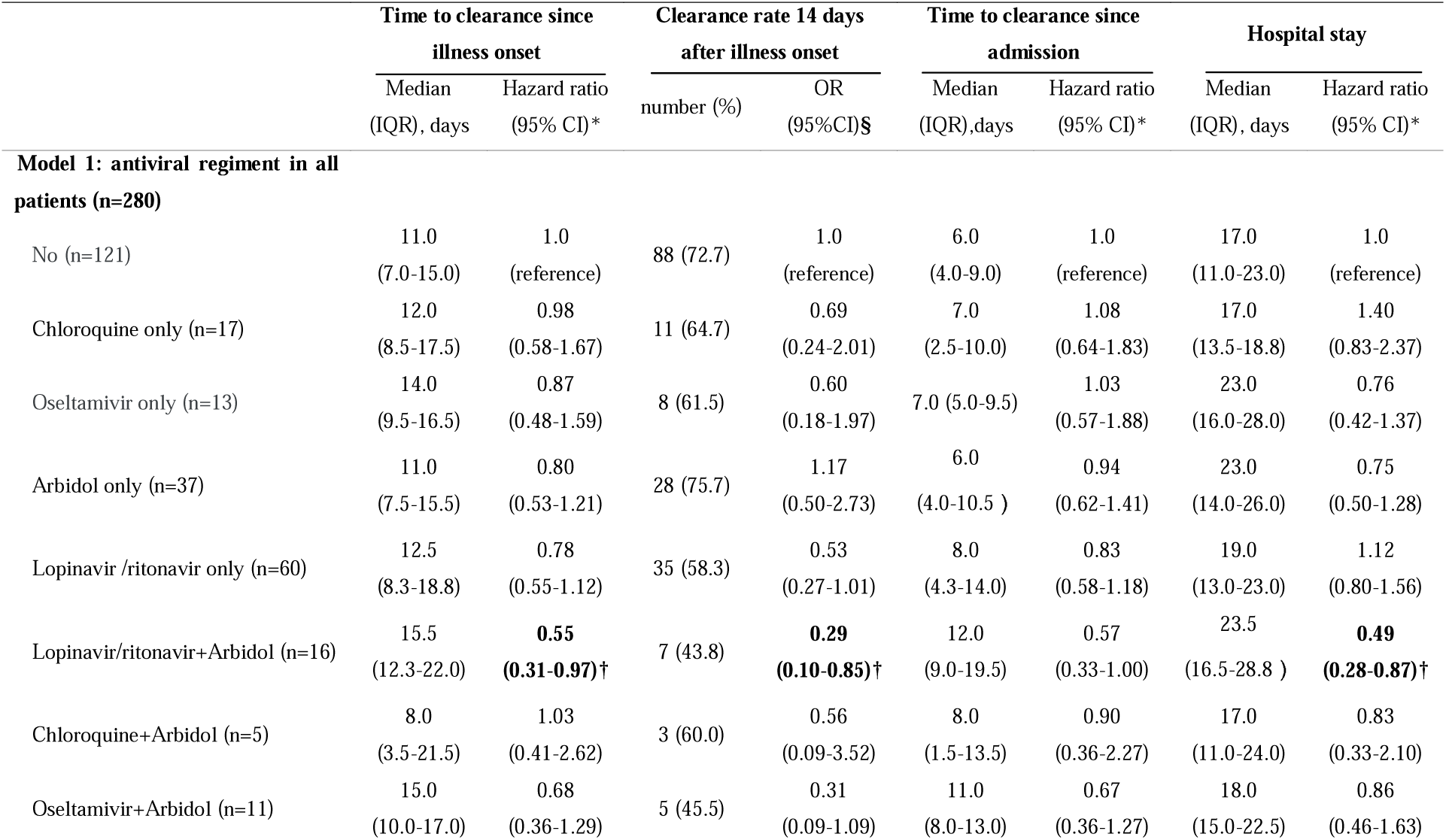
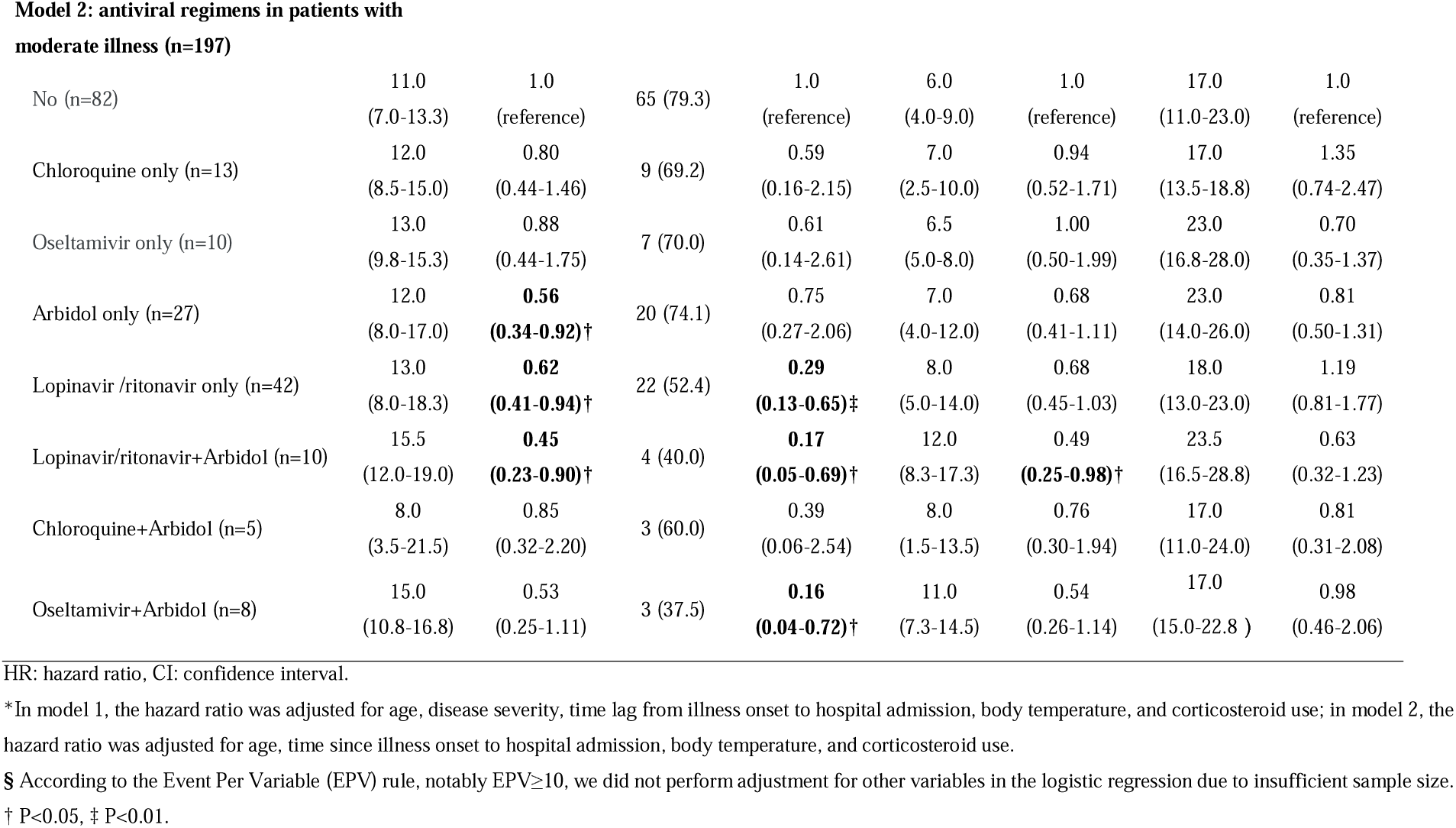
The associations between antiviral regimens and viral clearance in patients with COVID-19.

|  | Time to clearance since illness onset |  | Clearance rate 14 days after illness onset |  | Time to clearance since admission |  | Hospital stay |  |
| --- | --- | --- | --- | --- | --- | --- | --- | --- |
|  | Median (IQR), days | Hazard ratio (95% CI)* | number (%) | OR (95%CI)§ | Median (IQR),days | Hazard ratio (95% CI)* | Median (IQR), days | Hazard ratio (95% CI)* |
| **Model 1: antiviral regiment in all patients (n=280)** |  |  |  |  |  |  |  |  |
| No (n=121) | 11.0<br>(7.0-15.0) | 1.0<br>(reference) | 88 (72.7) | 1.0<br>(reference) | 6.0<br>(4.0-9.0) | 1.0<br>(reference) | 17.0<br>(11.0-23.0) | 1.0<br>(reference) |
| Chloroquine only (n=17) | 12.0<br>(8.5-17.5) | 0.98<br>(0.58-1.67) | 11 (64.7) | 0.69<br>(0.24-2.01) | 7.0<br>(2.5-10.0) | 1.08<br>(0.64-1.83) | 17.0<br>(13.5-18.8) | 1.40<br>(0.83-2.37) |
| Oseltamivir only (n=13) | 14.0<br>(9.5-16.5) | 0.87<br>(0.48-1.59) | 8 (61.5) | 0.60<br>(0.18-1.97) | 7.0 (5.0-9.5) | 1.03<br>(0.57-1.88) | 23.0<br>(16.0-28.0) | 0.76<br>(0.42-1.37) |
| Arbidol only (n=37) | 11.0<br>(7.5-15.5) | 0.80<br>(0.53-1.21) | 28 (75.7) | 1.17<br>(0.50-2.73) | 6.0<br>(4.0-10.5 ) | 0.94<br>(0.62-1.41) | 23.0<br>(14.0-26.0) | 0.75<br>(0.50-1.28) |
| Lopinavir /ritonavir only (n=60) | 12.5<br>(8.3-18.8) | 0.78<br>(0.55-1.12) | 35 (58.3) | 0.53<br>(0.27-1.01) | 8.0<br>(4.3-14.0) | 0.83<br>(0.58-1.18) | 19.0<br>(13.0-23.0) | 1.12<br>(0.80-1.56) |
| Lopinavir/ritonavir+Arbidol (n=16) | 15.5<br>(12.3-22.0) | **0.55**<br>**(0.31-0.97)†** | 7 (43.8) | **0.29**<br>**(0.10-0.85)†** | 12.0<br>(9.0-19.5) | 0.57<br>(0.33-1.00) | 23.5<br>(16.5-28.8 ) | **0.49**<br>**(0.28-0.87)†** |
| Chloroquine+Arbidol (n=5) | 8.0<br>(3.5-21.5) | 1.03<br>(0.41-2.62) | 3 (60.0) | 0.56<br>(0.09-3.52) | 8.0<br>(1.5-13.5) | 0.90<br>(0.36-2.27) | 17.0<br>(11.0-24.0) | 0.83<br>(0.33-2.10) |
| Oseltamivir+Arbidol (n=11) | 15.0<br>(10.0-17.0) | 0.68<br>(0.36-1.29) | 5 (45.5) | 0.31<br>(0.09-1.09) | 11.0<br>(8.0-13.0) | 0.67<br>(0.36-1.27) | 18.0<br>(15.0-22.5) | 0.86<br>(0.46-1.63) |
| **Model 2: antiviral regimens in patients with moderate illness (n=197)** |  |  |  |  |  |  |  |  |
| No (n=82) | 11.0<br>(7.0-13.3) | 1.0<br>(reference) | 65 (79.3) | 1.0<br>(reference) | 6.0<br>(4.0-9.0) | 1.0<br>(reference) | 17.0<br>(11.0-23.0) | 1.0<br>(reference) |
| Chloroquine only (n=13) | 12.0<br>(8.5-15.0) | 0.80<br>(0.44-1.46) | 9 (69.2) | 0.59<br>(0.16-2.15) | 7.0<br>(2.5-10.0) | 0.94<br>(0.52-1.71) | 17.0<br>(13.5-18.8) | 1.35<br>(0.74-2.47) |
| Oseltamivir only (n=10) | 13.0<br>(9.8-15.3) | 0.88<br>(0.44-1.75) | 7 (70.0) | 0.61<br>(0.14-2.61) | 6.5<br>(5.0-8.0) | 1.00<br>(0.50-1.99) | 23.0<br>(16.8-28.0) | 0.70<br>(0.35-1.37) |
| Arbidol only (n=27) | 12.0<br>(8.0-17.0) | **0.56**<br>**(0.34-0.92)†** | 20 (74.1) | 0.75<br>(0.27-2.06) | 7.0<br>(4.0-12.0) | 0.68<br>(0.41-1.11) | 23.0<br>(14.0-26.0) | 0.81<br>(0.50-1.31) |
| Lopinavir /ritonavir only (n=42) | 13.0<br>(8.0-18.3) | **0.62**<br>**(0.41-0.94)†** | 22 (52.4) | **0.29**<br>**(0.13-0.65)‡** | 8.0<br>(5.0-14.0) | 0.68<br>(0.45-1.03) | 18.0<br>(13.0-23.0) | 1.19<br>(0.81-1.77) |
| Lopinavir/ritonavir+Arbidol (n=10) | 15.5<br>(12.0-19.0) | **0.45**<br>**(0.23-0.90)†** | 4 (40.0) | **0.17**<br>**(0.05-0.69)†** | 12.0<br>(8.3-17.3) | 0.49<br>**(0.25-0.98)†** | 23.5<br>(16.5-28.8) | 0.63<br>(0.32-1.23) |
| Chloroquine+Arbidol (n=5) | 8.0<br>(3.5-21.5) | 0.85<br>(0.32-2.20) | 3 (60.0) | 0.39<br>(0.06-2.54) | 8.0<br>(1.5-13.5) | 0.76<br>(0.30-1.94) | 17.0<br>(11.0-24.0) | 0.81<br>(0.31-2.08) |
| Oseltamivir+Arbidol (n=8) | 15.0<br>(10.8-16.8) | 0.53<br>(0.25-1.11) | 3 (37.5) | **0.16**<br>**(0.04-0.72)†** | 11.0<br>(7.3-14.5) | 0.54<br>(0.26-1.14) | 17.0<br>(15.0-22.8 ) | 0.98<br>(0.46-2.06) |

HR: hazard ratio, CI: confidence interval.
\*In model 1, the hazard ratio was adjusted for age, disease severity, time lag from illness onset to hospital admission, body temperature, and corticosteroid use; in model 2, the hazard ratio was adjusted for age, time since illness onset to hospital admission, body temperature, and corticosteroid use.
§ According to the Event Per Variable (EPV) rule, notably $EPV \geq 10$ , we did not perform adjustment for other variables in the logistic regression due to insufficient sample size.
† $P < 0.05$ , ‡ $P < 0.01$ .

## Discussion

In this retrospective study, 89% of patients with COVID 19 achieved viral RNA clearance within 21 days. Time lag from illness onset to hospital admission, old age, disease severity, high body temperature, and corticosteroid use were associated with delayed clearance of viral RNA. The use of antiviral drugs (chloroquine, oseltamivir, arbidol, and lopinavir/ritonavir) had no benefit on such clearance. After adjusting for confounders, the use of lopinavir/ritonavir or the combination of two antiviral drugs tended to delay the clearance of viral RNA.

In this study, the median time from illness onset to viral RNA clearance was 12 days. This was compared to 15 days reported in patients from Wuhan [18], which might reflect the more severe nature of the latter patients. Currently, there is no consensus on the de-isolation management for patients with COVID-19, especially in asymptomatic subjects with high risk of infectivity. In this cohort, 89% of patients achieved viral RNA clearance within 21 days, and only 66% within 14 days. Specifically, all asymptomatic patients achieved viral RNA clearance within 21 days. These data suggested that isolation for at least 21 days after the onset of symptom or first positive PCR test is warranted in both patients without or with improving symptoms.

Symptomatic treatment and supportive care is the mainstay management of COVID-19. No antiviral drugs specific for COVID-19 have been approved, although there are ongoing clinical trials in COVID 19 evaluating the efficacy of various drugs approved for other conditions [19]. Chloroquine is an oral anti-malaria drug with in-vitro antiviral activity against SARS-CoV2 [20]. In some small-scale uncontrolled studies, chloroquine was reported to improve symptoms and clearance of viral RNA [15]. This drug is being recommended for the treatment of hospitalized COVID-19 patients in China and some countries [11, 17]. In this study, 28 patients were treated with chloroquine which did not appear to improve viral RNA clearance compared to supportive treatment. Large scale randomized controlled studies are needed to confirm its benefit, if any.

Lopinavir/ritonavir is a fixed dose combination of anti-HIV protease inhibitors acting against the viral 3CL protease [21]. Although it has been used to treat COVID-19 [22], its benefits has not been confirmed in a clinical trial involving COVID-19 patients with pneumonia in Wuhan [16]. In our study, apart from lack of benefit, the use of lopinavir/ ritonavir tended to delay viral RNA clearance, even after adjusting for factors including disease severity.

Arbidol is an antiviral drug approved in Russia and China for treating influenza. It acts against influenza A and B by inhibiting viral fusion with cellular membrane and blocking the virus entry into host cells [23]. In a retrospective cohort study, arbidol accelerated the clearance of viral RNA in patients with COVID-19 compared to empirical treatment [14]. In China, it has been recommended as one of the antiviral drugs for COVID-19 [11]. Oseltamivir is another drug approved for the treatment of influenza A and B [24], and has been used in some patients with COVID-19 [3, 7]. In this study, neither arbidol nor oseltamivir exhibited positive effects on the viral RNA clearance of COVID-19. Further, the combination of arbidol with another antiviral drug, oseltamivir or lopinavir/ritonavir, tended to delay clearance of viral RNA.

In this study, only 10% of patients received corticosteroid treatment compared to 50%-79% as reported in patients from Wuhan [7, 9]. Our analysis indicated that corticosteroid use was associated with delayed viral RNA clearance, in line with a recent report from Wuhan [18]. This may be due to selection bias since patients with serious/critical illness were more likely to be treated with corticosteroid. Moreover, corticosteroid can suppress the immune systemand delay clearance of viral RNA [25], calling for its cautious use in patients with COVID-19.

Older age and disease severity are major determinants of persistence of viral RNA. While the immune system weakens with aging [26], disease severity is a reflection of the host immunity. Indeed, over 35% of patients aged 65 years old or above had serious or critical illness. Here, public health measures such as social distancing, widespread testing, quarantine and contact tracing will not only reduce the spread of COVID-19 but also allows early detection of patients with mild disease for timely supportive treatment. This will help reduce the burden on the healthcare system for treating patients with serious illness and reduce morbidity mortality rates. In Guangzhou, with increased vigilance after the initial outbreak, these measures together with adequate medical facilities and support, the mortality rate was less than 1% compared to 3-4% during the early phase of the outbreak in Wuhan.

There were some limitations in this study. First, due to small sample size, our data regarding treatment efficacy of antiviral drugs should be interpreted with caution. Second, our patients had predominately mild to moderate illness and the results may not apply to those with serious or critical illness. Third, the RT-PCR test used to detect viral nuclear acid in this study may not be sensitive enough to detect samples with low viral load with possible false-negative results. Fourth, the viral RNA test was repeated every 2-3 days after hospitalization, therefore the time to viral RNA clearance may have a little bias. Lastly, it is a retrospective single-center study, and large-scale prospective studies are needed to confirm these findings.

In conclusion, in patients with COVID-19, a minimum of 21 days’ isolation since illness onset may be warranted even for asymptomatic patients. Old people are more likely to have serious illness with delayed clearance of viral RNA, and may require prolonged period of support and isolation. The use of antiviral drugs (chloroquine, oseltamivir, arbidol, and lopinavir/ritonavir) did not improve viral RNA clearance, while corticosteroid use was associated with delayed clearance of viral RNA and should be used with caution.

## Data Availability

No additional data available.

## Contributions

XC, YZ, BZ, and JW designed the study. XC, JZ, WH, XH, JC, HZ, SQ, and JW contributed to disease diagnosis and treatment and data collection. YZ and BZ analyzed data, XC, YZ, BZ, JCNC and JW wrote the paper. All authors reviewed the paper and approved the final version of the paper

## Acknowledgement

We thank all patients and care providers who participated in this study.

